# Admission Blood Pressure and Clinical Outcomes After Endovascular Treatment for Acute Basilar Artery Occlusion

**DOI:** 10.1101/2025.09.17.25336033

**Authors:** Xiao Dong, Yuanyuan Liu, Hongrui Ma, Baoying Song, Xuehong Chu, Wanwan Zhang, Yibing Guo, Erlan Yu, Xunming Ji, Chuanhui Li, Chuanjie Wu

**Affiliations:** Department of Neurology, Xuanwu Hospital, Capital Medical University, Beijing, China

**Author notes:** **Corresponding:** Prof. Chuanjie Wu, Postal Address: Xuanwu Hospital Capital Medical University, Beijing 100053, China. These authors contributed equally (Xiao Dong, Yuanyuan Liu).

**Keywords:** Blood Pressure, Basilar Artery Occlusion, Acute Ischemic Stroke, Endovascular Thrombectomy, Prognosis, Intercranial Hemorrhage

## Abstract

**BACKGROUND:** Basilar artery occlusion is a devastating subtype of stroke with low incidence. More than half of these patients fail to achieve favorable outcomes after endovascular treatment. Early blood pressure management may be an important determinant of prognosis; however, evidence for basilar artery occlusion remains limited and inconsistent.

**METHODS:** Patients with acute basilar artery occlusion who underwent endovascular treatment were screened using a prospective registry. The earliest non-invasive blood pressure measurements recorded in the emergency department were extracted. Associations between admission blood pressure and outcomes were analyzed using logistic regression for linear and restricted cubic splines for nonlinear trends. The primary outcome was favorable functional outcome at 90 days (modified Rankin Scale score ≤3).

**RESULTS:** A total of 387 patients with acute basilar artery occlusion treated with endovascular treatment were included in the analysis. Admission systolic and diastolic blood pressure showed inverted U-shaped associations with favorable functional outcomes, with thresholds of 154 and 97 mmHg, respectively. Relative to the threshold of 154 mmHg, each 10-mmHg increase in systolic blood pressure above the threshold was associated with reduced odds of a favorable outcome (adjusted OR, 0.71; 95% CI, 0.56-0.90), and each 10-mmHg decrease in systolic BP below the threshold was also associated with reduced odds of a favorable outcome (adjusted OR, 0.76; 95% CI, 0.58-0.99). Compared to the peak range (140–165 mmHg), systolic blood pressure both <140 mmHg and ≥165 mmHg were associated with lower odds of favorable outcomes (adjusted OR, 0.58; 95% CI, 0.35-0.95, adjusted OR, 0.66; 95% CI, 0.38-0.98, respectively). Higher blood pressure was linearly associated with a higher incidence of symptomatic intracranial hemorrhage.

**CONCLUSIONS:** In patients with acute basilar artery occlusion who underwent endovascular treatment, higher systolic and lower diastolic blood pressures were associated with a lower likelihood of favorable functional outcomes.

## INRODUCTION

Basilar artery occlusion (BAO) is a rare but life-threatening condition with high morbidity and mortality and accounts for approximately 1% of all acute ischemic strokes.^1,2^ Endovascular thrombectomy has proven effective in patients with BAO.^3,4^ A recent meta-analysis pooled several randomized trials comparing endovascular thrombectomy and standard medical therapy in patients with BAO, showing robust efficacy and safety of endovascular thrombectomy.^5^ However, nearly 55% of patients did not achieve a favorable functional outcome at 90 days (modified Rankin Scale [mRS] score ≤3) and 36% died within 90 days despite treatment with endovascular treatment (EVT).^5^ The early optimization of blood pressure (BP) may be a potential adjunctive strategy for improving outcomes in patients who undergo EVT. In acute ischemic stroke caused by large-vessel occlusion in the anterior circulation, evidence from several observational studies has indicated that BP at admission is related to clinical outcomes in a J- or U-shaped manner.^6–8^ A meta-analysis of data from seven randomized controlled trials also confirmed a nonlinear association with an inflection point at 140 mm Hg.^9^ However, related research on the posterior circulation is limited. One retrospective study suggested a linear relationship between BP at admission poor functional outcomes in patients with acute BAO.^10^ The study population showed a relatively low rate of successful reperfusion, partially because it included patients lacking adequate reperfusion, and high-quality evidence is still lacking in the current era of common reperfusion treatment. Given the limited evidence on BAO, this study aimed to assess the association between admission BP and clinical and imaging outcomes in patients with acute BAO patients who underwent EVT in a real-world setting.

## METHODS

### Study Population and Patient Selection

This cohort study used data from a prospective registry at Xuanwu Hospital, Capital Medical University (Beijing, China), which consecutively enrolled patients with ischemic stroke treated with EVT between August 2016 and December 2024.

All EVT procedures and treatments were administered according to current guidelines. The procedures were performed by neurointerventionalists using currently available, approved devices. At a 3-months follow-up, functional outcomes were assessed either during an on-site hospital visit, as recommended, or via telephone interviews conducted by trained neurologists. The EVT intervention process has been previously described.^11,12^

This study included patients from the host institution’s patient registry database, using the following inclusion criteria: (1) age ≥ 18 years; (2) confirmed occlusion of the basilar artery or V4 segments of bilateral vertebral arteries using magnetic resonance angiography, computed tomography angiography, or digital subtraction angiography; (3) treatment with EVT within 24 h of symptom onset or of the time the patient was known to be without symptoms, (4) a pretreatment mRS score ≤ 2, and (5) availability of admission BP parameters. The exclusion criteria were: (1) evidence of hemorrhage on admission computed tomography or magnetic resonance imaging before EVT and (2) occlusions involving both the anterior and posterior circulation arteries. This study complied with the STROBE checklist and was approved by the Ethics Committee of Xuanwu Hospital, Capital Medical University (No. [2017]030). All participants or their legal representatives provided written informed consent for study participation.

### Study Variables

We extracted the following variables from the patient database: demographic factors (sex, age), comorbidities (atrial fibrillation, hypertension, diabetes mellitus, prior cerebral ischemic/hemorrhagic stroke, pre-stroke mRS), admission BP parameters, baseline National Institute of Health Stroke Scale (NIHSS) scores, imaging parameters (lesion site, posterior circulation Alberta Stroke Program Early Computed Tomography Score [pc-ASPECTS], median Pons–Midbrain Index), stroke etiology, treatment details, and clinical outcomes. Admission BP, measured as systolic BP (SBP) and diastolic BP (DBP), was the earliest noninvasive measurement recorded during routine clinical assessments in the emergency department using an automated sphygmomanometer. Stroke etiology was classified according to the TOAST criteria (Trial of Org 10172 Acute Stroke Treatment). Occlusion sites were categorized as proximal, middle, or distal to the basilar artery and the V4 segment of the vertebral artery. pc-ASPECTS and median Pons–Midbrain Index scores were assessed using non-contrast enhanced computed tomography. Treatment details included administration of intravenous thrombolysis, anesthesia modality, time from symptom onset to admission, time from symptom onset to groin puncture, and time from symptom onset to successful reperfusion.

### Outcome Measures

The primary outcome was a favorable functional outcome, defined as an mRS score ≤3 at 90 days. The mRS is an ordinal measure, with scores ranging from 0 (no symptoms) to 6 (death). Secondary outcomes included functional independence (mRS ≤2) and excellent functional outcome (mRS ≤1) at 90 days, the ordinal mRS score at 90 days, successful reperfusion after endovascular thrombectomy (modified Thrombolysis in Cerebral Infarction score 2b or 3), and early neurological improvement (a decrease of at least 8 points in the NIHSS score from baseline or an NIHSS score of 0–2 at 24 h). Safety outcomes included symptomatic intracranial hemorrhage (sICH) within 24 h after EVT and all-cause mortality within 90 days. All efficacy and safety outcomes were assessed by qualified neurologists or trained staff.

### Statistical Analysis

Categorical variables were expressed as frequencies and percentages. Continuous variables were reported as mean ± standard deviation or as median and interquartile range, depending on the distribution. Missing values were replaced using multiple imputations. As we anticipated a J- or U-shaped association between admission BP and outcomes, we compared regression models with linear BP parameters against those with restricted cubic spline transformations using likelihood ratio tests. Restricted cubic splines were pre-specified with four knots placed at the 5th, 35th, 65th, and 95th percentiles of the BP distribution. When the transformation of the BP parameters did not improve the model fit, the linear model was chosen. We assessed whether SBP or DBP showed a stronger association with favorable functional outcomes at 90 day using Akaike Information Criterion.

When a J- or U-shaped nonlinear association between BP and the primary outcome was identified, we reported the threshold, defined as the BP point in time with the highest odds ratio (OR) of a favorable functional outcome, together with its 95% confidence interval (CI) obtained via bootstrap resampling. In the regression models, the threshold was used as the reference, and the risks associated with each 10-mmHg decrease below and each 10-mmHg increase above the threshold were estimated. For each outcome variable, multivariate binary logistic regression or ordinal logistic regression was used, as appropriate. All models were adjusted for age, sex, hypertension, baseline NIHSS score, baseline pc-ASPECTS, and the time from onset to admission. Ordinal logistic regression models were used to estimate the shift in the mRS score at 90 days after confirming the proportional odds assumption using the Brant test (p > 0.05). For interpretation, the scores were inverted so that the common odds ratio could be calculated for every one-point increase in mRS scores. In the sensitivity analysis, we examined the robustness and interpretability of the findings. First, when a J- or U-shaped nonlinear association in the primary outcome was present, we identified the BP values where the 95% CIs of the ORs crossed the reference line (OR = 1) in the model of primary outcome to facilitate categorical interpretation of the spline-derived relationships. These intersection points were used as cutoff values to classify the patients into three categories, with the interval between the intersection points defined as the peak range and designated as the reference group in the regression models. Second, we fitted multivariate models adjusted for an extended set of potential confounders, including age, sex, history of hypertension, diabetes mellitus, atrial fibrillation, previous stroke, pre-stroke mRS, baseline NIHSS, baseline pc-ASPECTS, occlusion site, etiology, intravenous thrombolysis, anesthesia modality, and onset-to-admission time. Third, we repeated the analysis using the complete case population. In the interaction analysis, we further assessed whether successful reperfusion modified the association between baseline SBP and favorable functional outcomes by introducing an interaction term into the multivariate logistic regression model.

R software (version 4.4.2) was used for all statistical analyses. Statistical significance was defined as a two-tailed p value < 0.05.

## RESULTS

### Patient Characteristics

Among 1496 patients who underwent EVT in the Xuanwu hospital within the defined study period, 387 met the inclusion and exclusion criteria and entered into the present analysis (Figure S1). The mean age was 62.7 years (SD 11.1); 77.7% (301/387) were male. The median NIHSS score was 20 (IQR, 13-33). The median time from symptom onset to admission was 417minutes (IQR, 263-613). Mean admission SBP was 155 mmHg (SD 23), while DBP was 86 mmHg (SD 15). The details were summarized in Table 1. The mRS score was available for 97.9% patients. 92 (46.2%) patients had favorable functional outcomes.

**Table 1.**
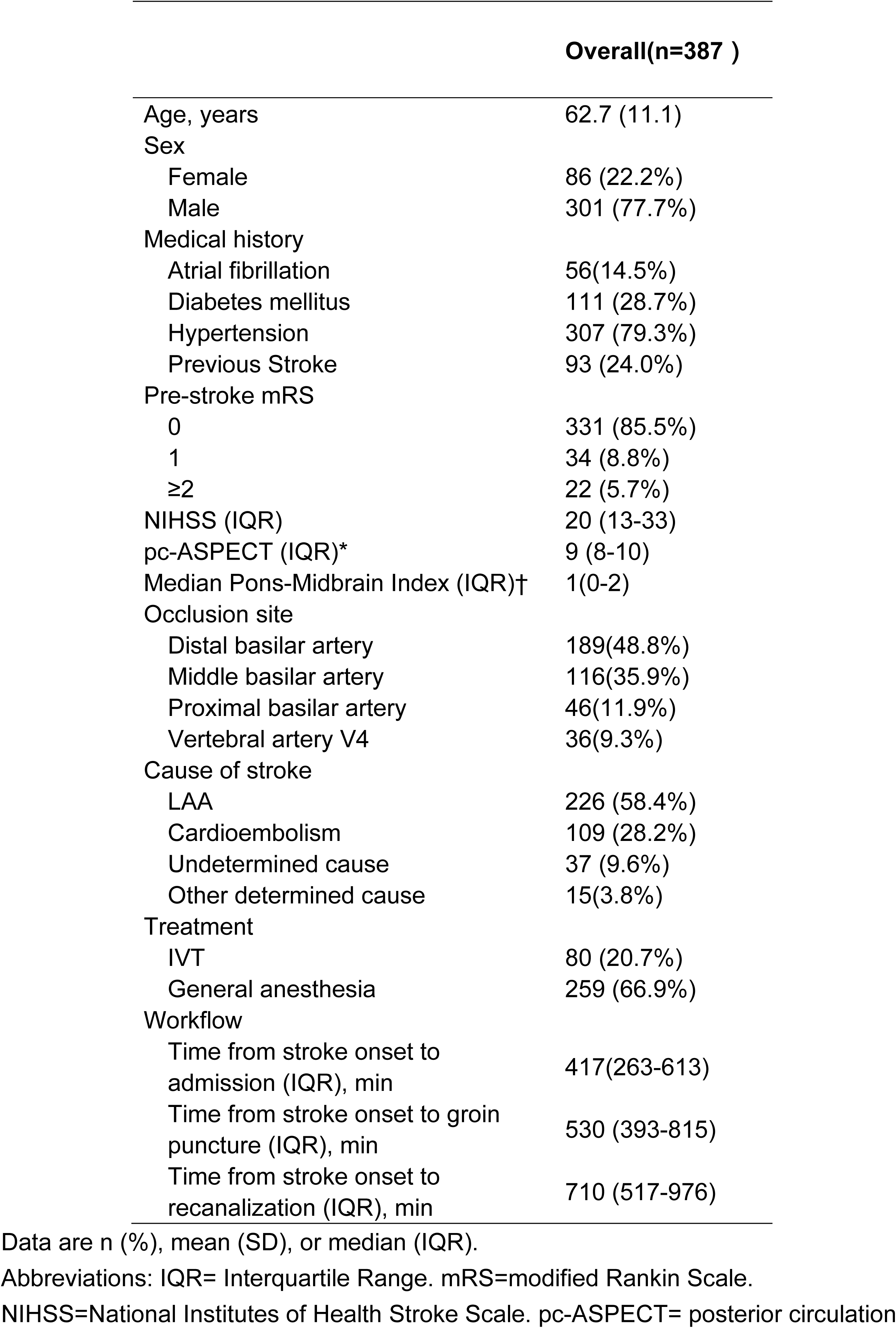
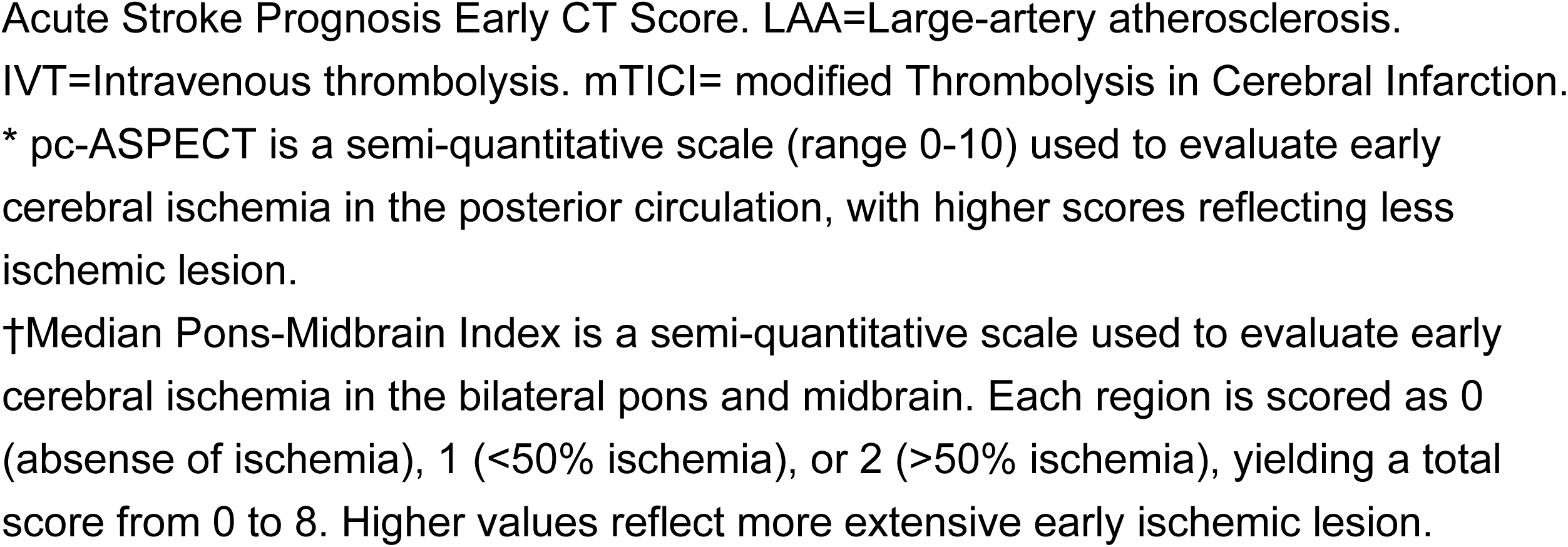
Characteristics of the Patients at Baseline.

### Primary Outcome

The association between admission SBP or DBP and favorable functional outcomes at 90 days was nonlinear (p for nonlinearity= 0.001 for SBP; p for nonlinearity= 0.013 for DBP). The goodness-of-fit of the model favored SBP (Akaike Information Criterion= 488) over DBP (Akaike Information Criterion=497). Using a restricted cubic spline, we observed an inverted U-shaped association between admission BP and favorable functional outcomes at 90 days, with thresholds of approximately 154 mmHg (95% CI, 145–159 mmHg) for SBP and 97 mmHg (95% CI 73–101 mmHg) for DBP (Figure 1). SBP was analyzed relative to a threshold of 154 mmHg. Each 10-mmHg increase in SBP above the threshold was associated with reduced odds of a favorable outcome (adjusted OR, 0.71; 95% CI, 0.56–0.90), whereas each 10-mmHg decrease in SBP below the threshold was also associated with reduced odds of a favorable outcome (adjusted OR, 0.76; 95% CI, 0.58–0.99).(Table 2) Similar patterns were observed for DBP, with increments above or below the threshold linked to lower odds of favorable outcomes (Figure S2).

**Figure 1.**
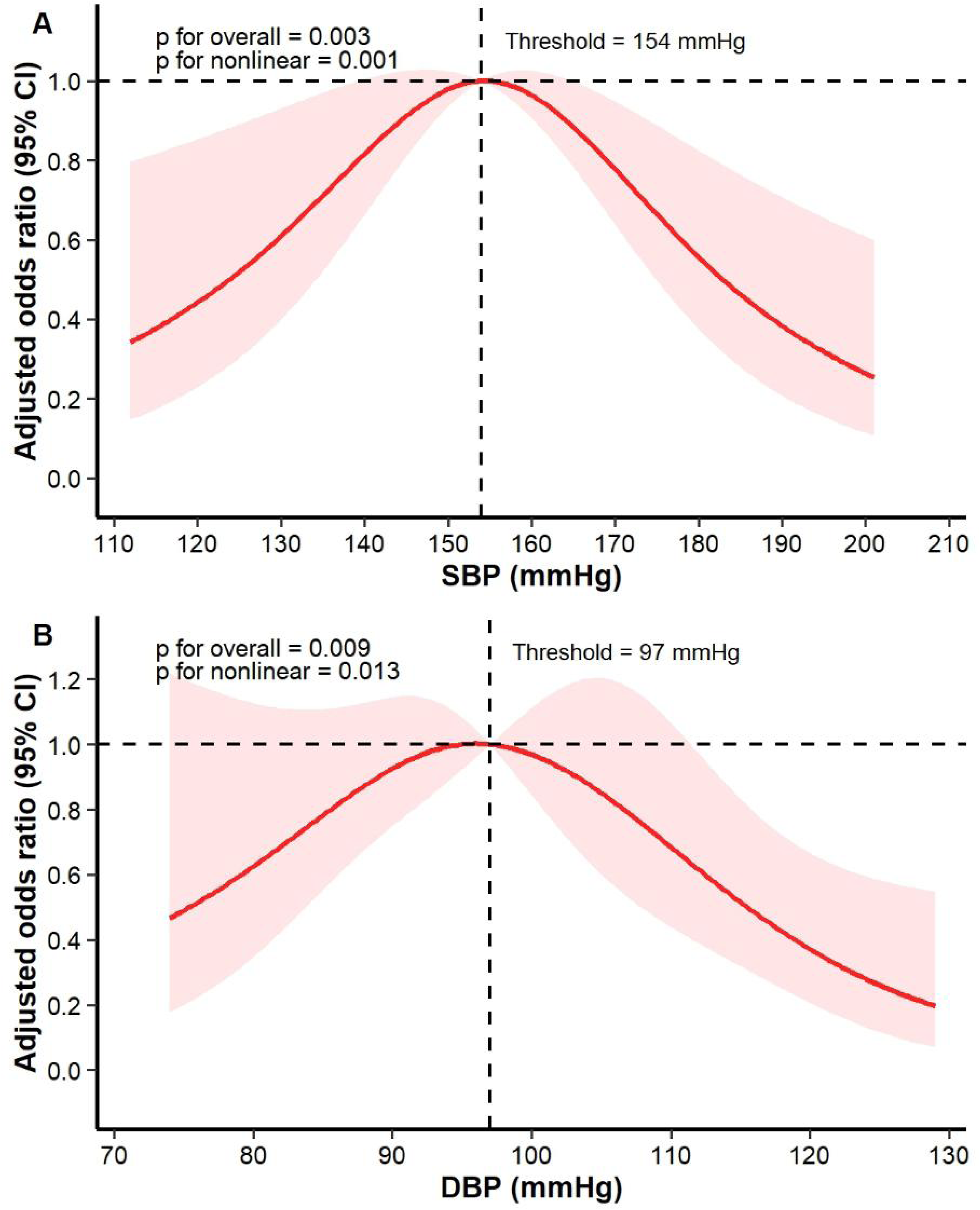
Restricted Cubic Spline Regression Models Representing the Relationship Between Admission BP and Favorable Outcome (mRS Scores 0-3) at 90 Days in Patients With BAO. (A) Relationship between admission SBP and favorable outcome (mRS scores 0-3); (B) Relationship between admission DBP and favorable outcome (mRS scores 0-3); Models were adjusted for age, sex, hypertension, baseline NIHSS, baseline pc-ASPECT, time from onset to admission. Red solid lines represent adjusted ORs, and light red shaded areas represent 95% CIs. Abbreviation: BAO = basilar artery occlusion; BP = blood pressure; mRS = modified Rankin Scale; OR = odds ratio; CI = confidence interval; SBP = systolic blood pressure; DBP = diastolic blood pressure.

**Table 2:**
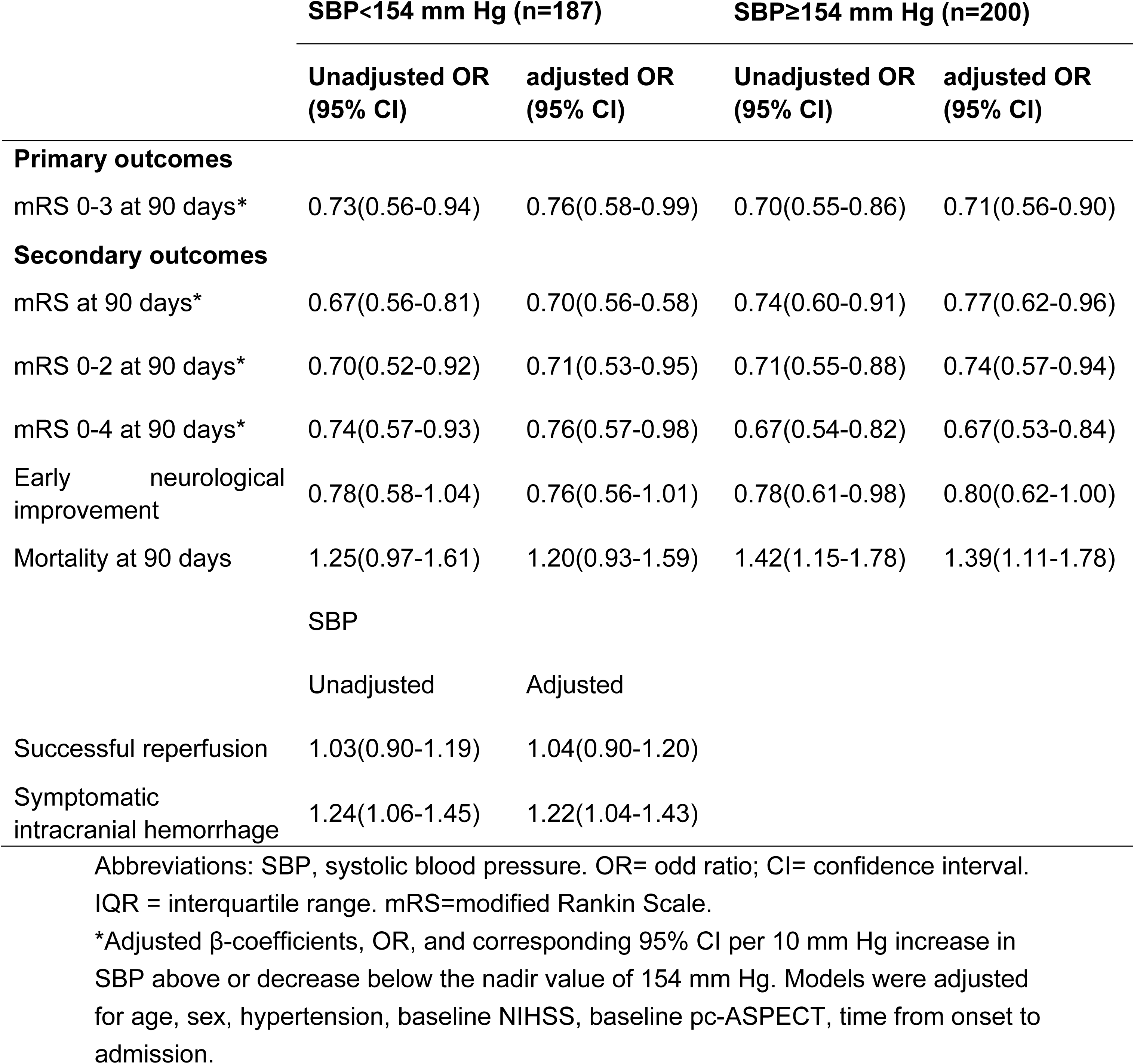
Association of Admission SBP With Outcomes in Univariable and Multivariable Analysis (at threshold).

### Secondary Outcome

A U-shaped association was also observed between admission SBP, 90-day mRS score and mortality, with the lowest ORs at 155 and 164 mmHg, respectively. An inverted U-shaped association was also observed between admission SBP and early neurological improvement, with the highest OR at 163 mmHg (Figure 2). When stratified at the spline-derived threshold from the primary outcome at 154 mmHg, both 10-mmHg decreases in SBP below the threshold and 10-mmHg increases in SBP above the threshold were linked to higher odds of worse functional outcomes. For mortality, an inverse pattern was observed in the unadjusted analysis. However, the association below the threshold was no longer significant after adjustment. For early neurological improvement, no significant associations were detected between SBP below or above 154 mmHg in the adjusted analyses. With respect to sICH, SBP showed a linear relationship; elevated SBP upon hospital admission was consistently associated with a greater probability of sICH (adjusted OR per 10-mmHg increase, 1.22; 95% CI, 1.04–1.43; Table 2, Figure 2).

**Figure 2.**
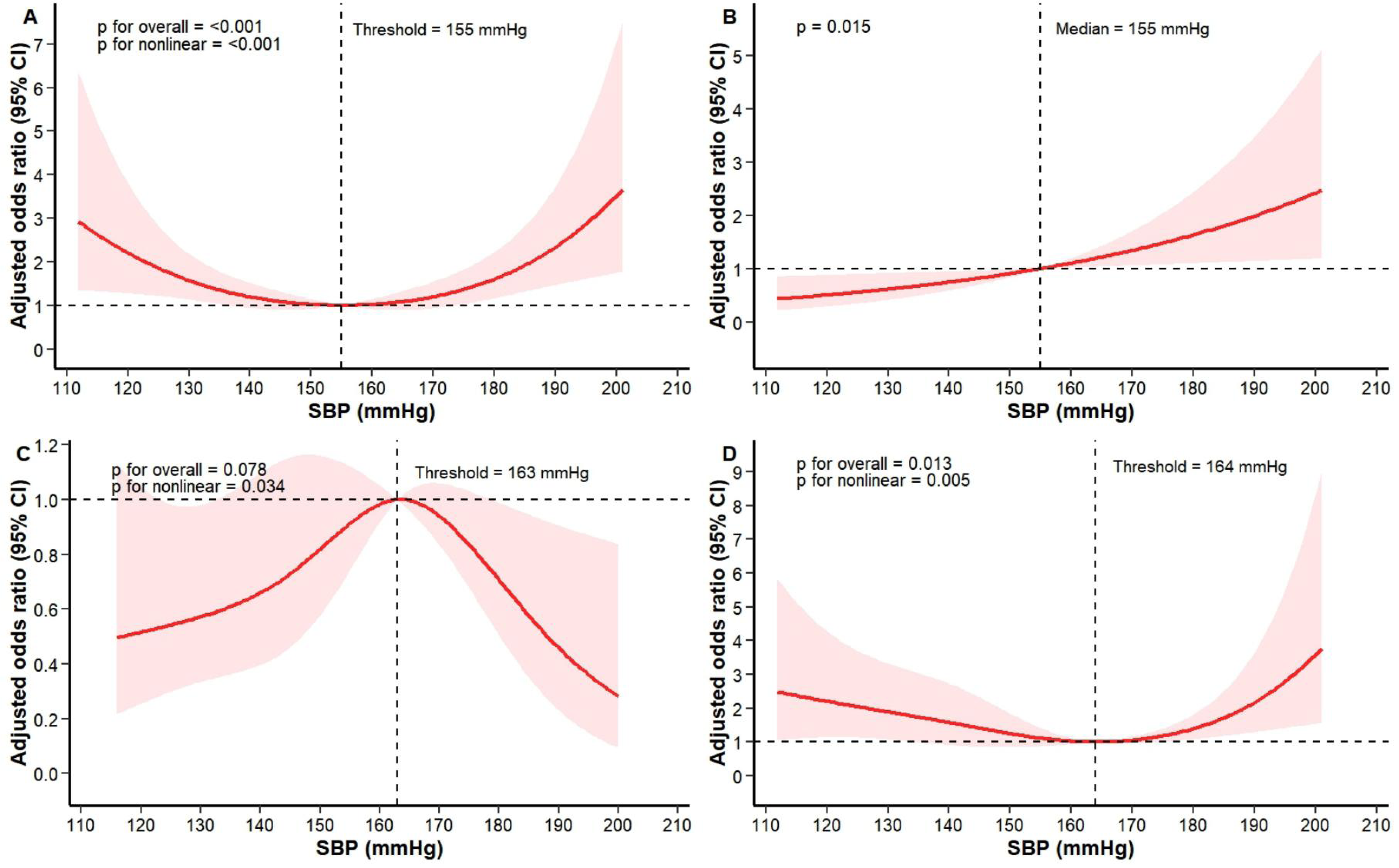
Regression Models Representing the Relationship Between Admission SBP and mRS score at 90 d, sICH, ENI, and 90-day mortality in Patients With BAO. (A) Relationship between admission SBP and mRS score at 90 d (restricted cubic spline model); (B) Relationship between admission SBP and sICH (linear model); (C)Relationship between admission SBP and ENI (restricted cubic spline model); (D) Relationship between admission SBP and 90-day mortality (restricted cubic spline model). Models were adjusted for age, sex, hypertension, baseline NIHSS, baseline pc-ASPECT, time from onset to admission. Red solid lines represent adjusted ORs, and light red shaded areas represent 95% CIs. Abbreviation: BAO = basilar artery occlusion; mRS = modified Rankin Scale; SBP = systolic blood pressure; ENI= early neurological improvement; sICH= symptomatic intracranial hemorrhage; OR = odds ratio; CI = confidence interval.

### Sensitivity Analysis

On the restricted cubic spline curve of SBP with the primary outcome, the two intersection points where the 95% CIs of the ORs crossed the reference line (OR = 1) were located at 140 and 165 mmHg. Based on these cutoffs, the study population was classified into three groups: SBP <140 mmHg, SBP = 140-165 mmHg (peak range), and SBP ≥165 mmHg. Statistically significant differences were observed in the proportion of patients with hypertension, pre-stroke mRS score, stroke etiology, and occlusion site among the three groups. The detailed baseline characteristics of the three groups are presented in Table S1. Favorable functional outcomes at 90 days were achieved in 41.7%, 55.0%, and 39.8% of the patients in these groups, respectively (Figure S3). Compared with patients in the peak range, those with SBP <140 mmHg or ≥165 mmHg had reduced odds of favorable outcomes in the adjusted model (SBP <140 mmHg: adjusted OR, 0.66; 95% CI, 0.38–0.98; SBP ≥165 mmHg: adjusted OR, 0.58; 95% CI, 0.35–0.95; Table 3). The inverted U-shaped association persisted after adjusting for extended confounders and complete-case analyses (Figure S4, Figure S5). No evidence was found indicating that reperfusion status influenced the association between SBP and favorable functional outcomes (P for interaction = 0.630).

**Table 3.**
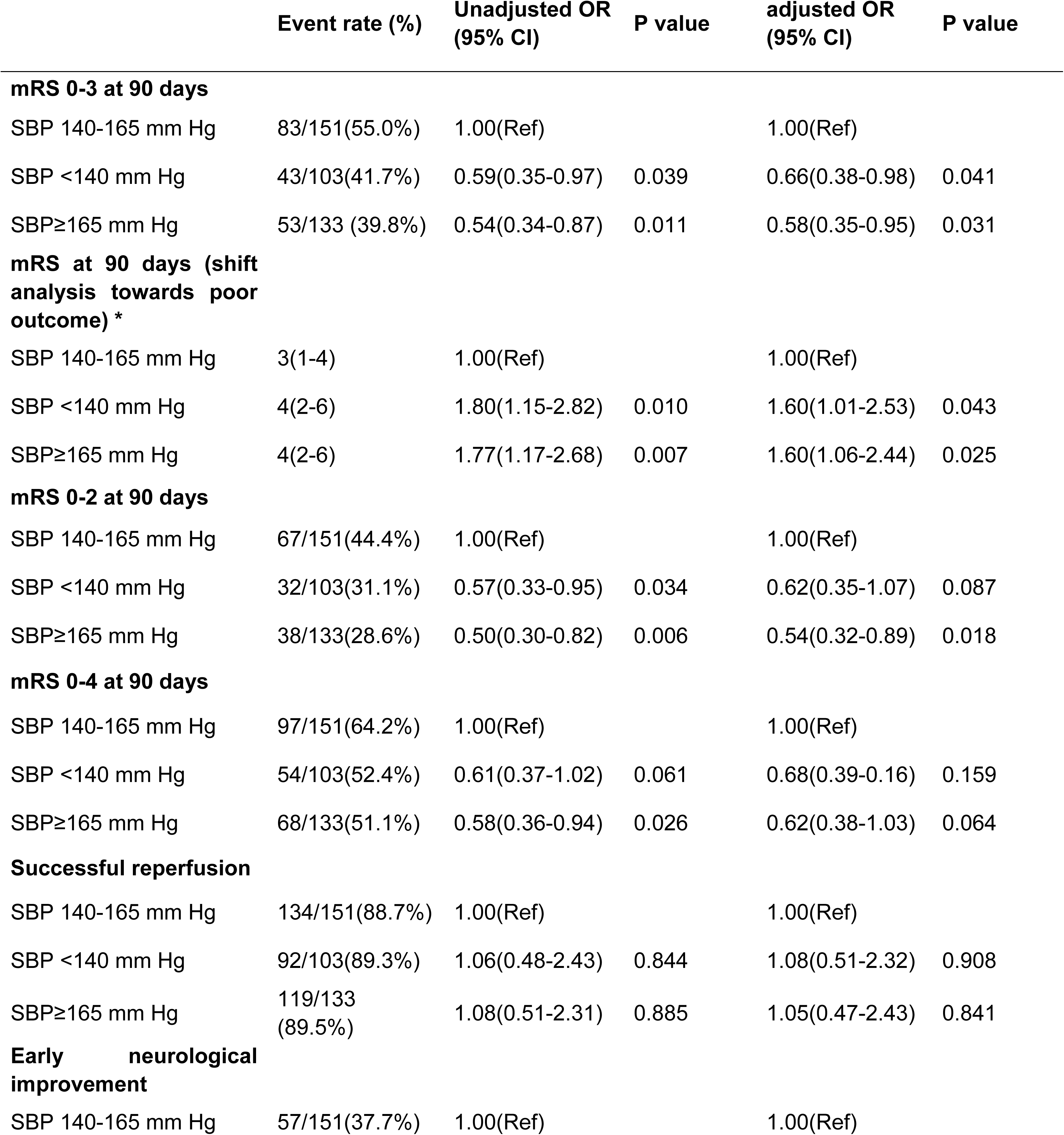
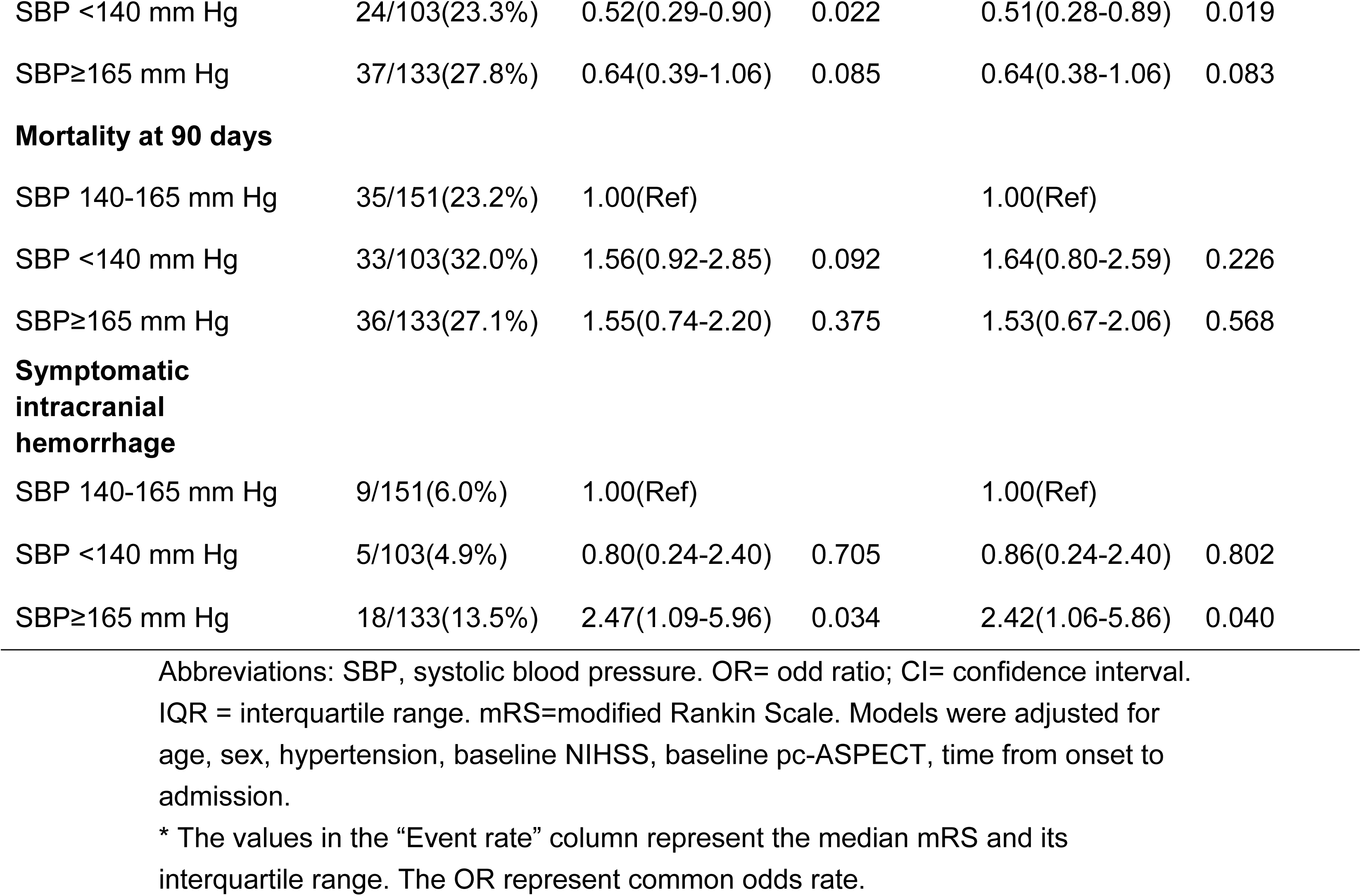
Association of Admission SBP With Outcomes in Univariable and Multivariable Analysis.

|  | Event rate (%) | Unadjusted OR<br>(95% CI) | P value | adjusted OR<br>(95% CI) | P value |
| --- | --- | --- | --- | --- | --- |
| **mRS 0-3 at 90 days** |  |  |  |  |  |
| SBP 140-165 mm Hg | 83/151(55.0%) | 1.00(Ref) |  | 1.00(Ref) |  |
| SBP <140 mm Hg | 43/103(41.7%) | 0.59(0.35-0.97) | 0.039 | 0.66(0.38-0.98) | 0.041 |
| SBP≥165 mm Hg | 53/133 (39.8%) | 0.54(0.34-0.87) | 0.011 | 0.58(0.35-0.95) | 0.031 |
| **mRS at 90 days (shift analysis towards poor outcome) \*** |  |  |  |  |  |
| SBP 140-165 mm Hg | 3(1-4) | 1.00(Ref) |  | 1.00(Ref) |  |
| SBP <140 mm Hg | 4(2-6) | 1.80(1.15-2.82) | 0.010 | 1.60(1.01-2.53) | 0.043 |
| SBP≥165 mm Hg | 4(2-6) | 1.77(1.17-2.68) | 0.007 | 1.60(1.06-2.44) | 0.025 |
| **mRS 0-2 at 90 days** |  |  |  |  |  |
| SBP 140-165 mm Hg | 67/151(44.4%) | 1.00(Ref) |  | 1.00(Ref) |  |
| SBP <140 mm Hg | 32/103(31.1%) | 0.57(0.33-0.95) | 0.034 | 0.62(0.35-1.07) | 0.087 |
| SBP≥165 mm Hg | 38/133(28.6%) | 0.50(0.30-0.82) | 0.006 | 0.54(0.32-0.89) | 0.018 |
| **mRS 0-4 at 90 days** |  |  |  |  |  |
| SBP 140-165 mm Hg | 97/151(64.2%) | 1.00(Ref) |  | 1.00(Ref) |  |
| SBP <140 mm Hg | 54/103(52.4%) | 0.61(0.37-1.02) | 0.061 | 0.68(0.39-1.16) | 0.159 |
| SBP≥165 mm Hg | 68/133(51.1%) | 0.58(0.36-0.94) | 0.026 | 0.62(0.38-1.03) | 0.064 |
| **Successful reperfusion** |  |  |  |  |  |
| SBP 140-165 mm Hg | 134/151(88.7%) | 1.00(Ref) |  | 1.00(Ref) |  |
| SBP <140 mm Hg | 92/103(89.3%) | 1.06(0.48-2.43) | 0.844 | 1.08(0.51-2.32) | 0.908 |
| SBP≥165 mm Hg | 119/133<br>(89.5%) | 1.08(0.51-2.31) | 0.885 | 1.05(0.47-2.43) | 0.841 |
| **Early neurological improvement** |  |  |  |  |  |
| SBP 140-165 mm Hg | 57/151(37.7%) | 1.00(Ref) |  | 1.00(Ref) |  |
| SBP <140 mm Hg | 24/103(23.3%) | 0.52(0.29-0.90) | 0.022 | 0.51(0.28-0.89) | 0.019 |
| SBP≥165 mm Hg | 37/133(27.8%) | 0.64(0.39-1.06) | 0.085 | 0.64(0.38-1.06) | 0.083 |
| **Mortality at 90 days** |  |  |  |  |  |
| SBP 140-165 mm Hg | 35/151(23.2%) | 1.00(Ref) |  | 1.00(Ref) |  |
| SBP <140 mm Hg | 33/103(32.0%) | 1.56(0.92-2.85) | 0.092 | 1.64(0.80-2.59) | 0.226 |
| SBP≥165 mm Hg | 36/133(27.1%) | 1.55(0.74-2.20) | 0.375 | 1.53(0.67-2.06) | 0.568 |
| **Symptomatic intracranial hemorrhage** |  |  |  |  |  |
| SBP 140-165 mm Hg | 9/151(6.0%) | 1.00(Ref) |  | 1.00(Ref) |  |
| SBP <140 mm Hg | 5/103(4.9%) | 0.80(0.24-2.40) | 0.705 | 0.86(0.24-2.40) | 0.802 |
| SBP≥165 mm Hg | 18/133(13.5%) | 2.47(1.09-5.96) | 0.034 | 2.42(1.06-5.86) | 0.040 |

Abbreviations: SBP, systolic blood pressure. OR= odd ratio; CI= confidence interval.
IQR = interquartile range. mRS=modified Rankin Scale. Models were adjusted for age, sex, hypertension, baseline NIHSS, baseline pc-ASPECT, time from onset to admission.
\* The values in the “Event rate” column represent the median mRS and its interquartile range. The OR represent common odds rate.

## DISCUSSION

This prospective cohort study included 387 patients treated with EVT for acute ischemic stroke caused by BAO in whom the admission SBP and DBP exhibited inverted U-shaped associations with favorable functional outcomes at 90 days and thresholds at 154 and 97 mmHg. Deviations from these thresholds in either direction– higher or lower BP–were associated with lower odds of favorable functional outcomes. Regarding safety outcomes, SBP showed a linear association with sICH, and higher SBP and DBP were associated with an increased risk of sICH.

Our findings are consistent with those of previous studies on acute ischemic stroke and anterior circulation EVT, where admission BP has frequently shown a J- or U-shaped association with clinical outcomes. However, the optimal SBP varies substantially across studies. In a cohort of 3,180 patients with anterior circulation stroke treated with EVT, both J- and U-shaped relationships were observed between SBP and poor functional outcomes at 90 d, mortality, and NIHSS at 24–48 hours, with SBP >150 mmHg predicting a worse prognosis.^6^ A post-hoc analysis of a randomized controlled trial on anterior circulation thrombectomy reported a U-shaped association between admission BP and poor functional outcomes, with a nadir of approximately 120 mmHg.^7^ Another study found that patients whose BP fell within the pre-specified normotensive SBP range of 155–220 mmHg had significantly lower 90-d mortality.^13^ These inconsistencies may be explained by variations in the inclusion criteria and baseline characteristics of the study population.

Currently, there is insufficient evidence regarding the association between BP at admission and clinical outcomes in patients with posterior circulation stroke. In a cohort of 826 patients diagnosed with BAO who underwent reperfusion therapy from the BASILAR registry, higher admission SBP and DBP were linearly associated with poorer functional outcomes, unlike the inverted U-shaped association we observed. ^10^Several factors may explain this discrepancy. First, treatment heterogeneity and lower reperfusion success in the previous study could mask the nonlinearity. A previous study enrolled individuals who received intravenous thrombolysis or endovascular treatment, with a successful reperfusion rate of only 64.3%, whereas the rate in the present cohort was approximately 90%. Second, hypertension was more prevalent in the present cohort, which may have skewed the cerebral autoregulation curve to the right, thereby amplifying the adverse effects of low BP. Finally, our statistical methods explicitly accommodated nonlinear relationships, whereas the previous study used a linear model, potentially obscuring nonlinear relationships.

The relationship among successful reperfusion, admission BP, and functional outcomes remains unclear. In the present study, BP at admission was not associated with successful reperfusion, and no evidence indicated that the reperfusion status influenced the link between SBP and functional outcomes. In a study based on the BASILAR registry, admission SBP >140 mmHg was associated with a lower likelihood of successful reperfusion than SBP <140 mmHg was, but the relationship between SBP and functional outcomes remained unchanged across reperfusion statuses.^10^ In contrast, research on anterior circulation occlusion found that the relationship between BP and prognosis differs across reperfusion statuses.^14–16^ In addition, we found a significant association between elevated BP and an increased occurrence of sICH. The BASILAR registry showed a similar trend, but it did not reach statistical significance.^10^

Several mechanisms may explain the inverted U-shaped association seen between admission BP and favorable functional outcomes in EVT. In large-vessel occlusion, ischemia and hypoxia can transiently impair autoregulation, rendering cerebral blood flow more vulnerable to fluctuations in BP. On the right side of the inverted U-shaped curve, a higher admission BP together with reperfusion injury after EVT may aggravate blood–brain barrier disruption, thereby increasing the early post-stroke risks of cerebral hemorrhage and edema. ^17–21^ Consistent with this mechanism, the present study found that a higher admission SBP was significantly associated with a greater incidence of early sICH. Another possible explanation is that a higher BP may indicate larger infarct volumes, suboptimal chronic hypertension control, or poor collateral circulation status, all of which are associated with worse clinical outcomes.^22,23^ However, because data on these variables were unavailable in our cohort, we could not directly test these hypotheses. On the left side of the inverted

U-shaped association, a lower BP may indicate compromised perfusion of the ischemic penumbra and a larger infarct. In contrast, hypotension may reflect a poor systemic status (e.g., hypovolemia or cardiac dysfunction). Specific features of the posterior circulation may amplify these adverse effects. First, the vertebrobasilar system supplies the medulla oblongata, the primary site of cardiorespiratory reflex integration; thus, BP abnormalities may directly precipitate life-threatening complications such as respiratory failure and circulatory instability. ^24^ Second, previous studies have shown that autoregulation in the posterior circulatory artery is less efficient than that in the anterior circulatory artery, which may be related to relatively sparse collaterals in the vertebrobasilar territory.^25,26^

Current American Stroke Association and European Stroke Association guidelines recommend aiming for a BP <185/110 mmHg before EVT.^27,28^ However, high-quality evidence supporting this threshold is limited, and the optimal target remains uncertain. A meta-analysis of seven randomized controlled trials (n=1753) showed that the admission SBP (<140 vs ≥ 140 mmHg) did not significantly affect the efficacy of EVT in anterior circulation, suggesting that SBP at presentation should not be considered a reason to defer or delay the procedure.^9^ But these results should be further investigated in randomized controlled trials. Owing to its unique anatomical and physiological features, BP management in BAO may differ from that in anterior circulation occlusions. These patients have been largely overlooked in previous randomized controlled trials of post-thrombectomy BP management. Our study provides a potential SBP range of 140–165 that may guide the design of SBP-targeting trials in patients with acute BAO who have undergone EVT. ^29–32^ Therefore, RCT trials are needed to determine the optimal pre-EVT BP targets, explicitly including posterior circulation strokes, to address the current gap in evidence.

Some limitations of this study deserve attention. First, given the observational design of the study, the associations cannot be interpreted as causal. Second, the single-center nature of this cohort may have restricted the generalizability of our results to a broader population. Third, information on antihypertensive treatment after hospital admission but before EVT initiation was unavailable, limiting further assessment of the relationship between preprocedural EVT BP and outcomes. Finally, the lack of records on the collateral status and infarct volume may have introduced unaddressed confounders into our model.

## CONCLUSIONS

Admission SBP and DBP demonstrated inverted U-shaped associations with favorable functional outcomes in patients with acute BAO who underwent EVT, and deviations from the thresholds (SBP = 154 mmHg, DBP = 97 mmHg) were associated with a lower likelihood of favorable outcomes. An admission SBP between 140 mmHg and 165 mmHg was associated with the highest likelihood of favorable functional outcomes. These data provide pragmatic insights to guide pre-EVT BP management. Further validation through multicenter randomized trials is warranted.

## Data Availability

All relevant data are available from the corresponding author upon reasonable request.

## Non-standard Abbreviations and Acronyms

BAO: basilar artery occlusion
EVT: endovascular treatment
pc-ASPECTS: posterior circulation Alberta Stroke Program Early Computed Tomography Score
sICH: symptomatic intracranial hemorrhage

## Acknowledgments

None

## Source of funding

This trial was funded by National Natural Science Foundation of China (grant number 82271507), Beijing Natural Science Foundation (grant number JQ24041), Noncommunicable Chronic Diseases - National Science and Technology Major Project (grant number 2023ZD0505403).

## Disclosures

None

## SUPPLEMENTAL MATERIAL

Table S1

Figures S1–S5

## Notes

### Competing Interest Statement

The authors have declared no competing interest.

### Clinical Trial

Not applicable. This study does not involve a clinical trial or prospective interventional study

### Author Declarations

This study was approved by the Ethics Committee of Xuanwu Hospital, Capital Medical University (No. [2017]030). All participants or their legal representatives provided written informed consent for study participation.

## References

1. Alemseged F, Nguyen TN, Alverne FM, Liu X, Schonewille WJ, Nogueira RG. Endovascular Therapy for Basilar Artery Occlusion. Stroke. 2023;54:1127–1137. doi: 10.1161/strokeaha.122.040807

2. Lindsberg PJ, Pekkola J, Strbian D, Sairanen T, Mattle HP, Schroth G. Time window for recanalization in basilar artery occlusion: Speculative synthesis. Neurology. 2015;85:1806–1815. doi: 10.1212/wnl.0000000000002129

3. Jovin TG, Li C, Wu L, Wu C, Chen J, Jiang C, Shi Z, Gao Z, Song C, Chen W, et al. Trial of Thrombectomy 6 to 24 Hours after Stroke Due to Basilar-Artery Occlusion. N Engl J Med. 2022;387:1373–1384. doi: 10.1056/NEJMoa2207576

4. Tao C, Nogueira RG, Zhu Y, Sun J, Han H, Yuan G, Wen C, Zhou P, Chen W, Zeng G, et al. Trial of Endovascular Treatment of Acute Basilar-Artery Occlusion. N Engl J Med. 2022;387:1361–1372. doi: 10.1056/NEJMoa2206317

5. Nogueira RG, Jovin TG, Liu X, Hu W, Langezaal LCM, Li C, Dai Q, Tao C, Mont’Alverne FJA, Ji X, et al. Endovascular therapy for acute vertebrobasilar occlusion (VERITAS): a systematic review and individual patient data meta-analysis. Lancet. 2025;405:61–69. doi: 10.1016/s0140-6736(24)01820-8

6. van den Berg SA, Uniken Venema SM, Mulder M, Treurniet KM, Samuels N, Lingsma HF, Goldhoorn RB, Jansen IGH, Coutinho JM, Roozenbeek B, et al. Admission Blood Pressure in Relation to Clinical Outcomes and Successful Reperfusion After Endovascular Stroke Treatment. Stroke. 2020;51:3205–3214. doi: 10.1161/strokeaha.120.029907

7. Mulder M, Ergezen S, Lingsma HF, Berkhemer OA, Fransen PSS, Beumer D, van den Berg LA, Lycklama À Nijeholt G, Emmer BJ, van der Worp HB, et al. Baseline Blood Pressure Effect on the Benefit and Safety of Intra-Arterial Treatment in MR CLEAN (Multicenter Randomized Clinical Trial of Endovascular Treatment of Acute Ischemic Stroke in the Netherlands). Stroke. 2017;48:1869–1876. doi: 10.1161/strokeaha.116.016225

8. Mortality and Disability According to Baseline Blood Pressure in Acute Ischemic Stroke Patients Treated by Thrombectomy: A Collaborative Pooled Analysis. J Am Heart Assoc. 2017;6:e004193. doi: 10.1161/jaha.117.004193

9. Samuels N, van de Graaf RA, Mulder M, Brown S, Roozenbeek B, van Doormaal PJ, Goyal M, Campbell BCV, Muir KW, Agrinier N, et al. Admission systolic blood pressure and effect of endovascular treatment in patients with ischaemic stroke: an individual patient data meta-analysis. Lancet Neurol. 2023;22:312–319. doi: 10.1016/s1474-4422(23)00076-5

10. Cao Y, Li R, Jiang S, Guo J, Luo X, Miao J, Liu J, Zheng B, Du J, Zhang Y, et al. The Relationship Between Admission Blood Pressure and Clinical Outcomes for Acute Basilar Artery Occlusion. Front Neurosci. 2022;16:900868. doi: 10.3389/fnins.2022.900868

11. Zhao W, Shang S, Li C, Wu L, Wu C, Chen J, Song H, Zhang H, Zhang Y, Duan J, et al. Long-term outcomes of acute ischemic stroke patients treated with endovascular thrombectomy: A real-world experience. J Neurol Sci. 2018;390:77–83. doi: 10.1016/j.jns.2018.03.004

12. Zhao W, Che R, Shang S, Wu C, Li C, Wu L, Chen J, Duan J, Song H, Zhang H, et al. Low-Dose Tirofiban Improves Functional Outcome in Acute Ischemic Stroke Patients Treated With Endovascular Thrombectomy. Stroke. 2017;48:3289–3294. doi: 10.1161/strokeaha.117.019193

13. Stead LG, Gilmore RM, Decker WW, Weaver AL, Brown RD, Jr. Initial emergency department blood pressure as predictor of survival after acute ischemic stroke. Neurology. 2005;65:1179–1183. doi: 10.1212/01.wnl.0000180939.24845.22

14. Martins AI, Sargento-Freitas J, Silva F, Jesus-Ribeiro J, Correia I, Gomes JP, Aguiar-Gonçalves M, Cardoso L, Machado C, Rodrigues B, et al. Recanalization Modulates Association Between Blood Pressure and Functional Outcome in Acute Ischemic Stroke. Stroke. 2016;47:1571–1576. doi: 10.1161/strokeaha.115.012544

15. Hong L, Cheng X, Lin L, Bivard A, Ling Y, Butcher K, Dong Q, Parsons M. The blood pressure paradox in acute ischemic stroke. Ann Neurol. 2019;85:331–339. doi: 10.1002/ana.25428

16. Kim BJ, Singh N, Kim H, Menon BK, Almekhlafi M, Ryu WS, Kim JT, Kang J, Baik SH, Kim JY, et al. Association between blood pressure and endovascular treatment outcomes differs by baseline perfusion and reperfusion status. Sci Rep. 2023;13:13776. doi: 10.1038/s41598-023-40572-0

17. Horsch AD, Dankbaar JW, van Seeters T, Niesten JM, Luitse MJ, Vos PC, van der Schaaf IC, Biessels GJ, van der Graaf Y, Kappelle LJ, et al. Relation between stroke severity, patient characteristics and CT-perfusion derived blood-brain barrier permeability measurements in acute ischemic stroke. Clin Neuroradiol. 2016;26:415–421. doi: 10.1007/s00062-015-0375-1

18. Bang OY, Saver JL, Alger JR, Shah SH, Buck BH, Starkman S, Ovbiagele B, Liebeskind DS. Patterns and predictors of blood-brain barrier permeability derangements in acute ischemic stroke. Stroke. 2009;40:454–461. doi: 10.1161/strokeaha.108.522847

19. van den Kerkhof M, de Jong JJA, Voorter PHM, Postma AA, Kroon AA, van Oostenbrugge RJ, Jansen JFA, Backes WH. Blood-Brain Barrier Integrity Decreases With Higher Blood Pressure: A 7T DCE-MRI Study. Hypertension. 2024;81:2162–2172. doi: 10.1161/hypertensionaha.123.22617

20. Desilles JP, Rouchaud A, Labreuche J, Meseguer E, Laissy JP, Serfaty JM, Lapergue B, Klein IF, Guidoux C, Cabrejo L, et al. Blood-brain barrier disruption is associated with increased mortality after endovascular therapy. Neurology. 2013;80:844–851. doi: 10.1212/WNL.0b013e31828406de

21. Mohammadi MT, Dehghani GA. Acute hypertension induces brain injury and blood-brain barrier disruption through reduction of claudins mRNA expression in rat. Pathol Res Pract. 2014;210:985–990. doi: 10.1016/j.prp.2014.05.007

22. Wartenberg KE, Mayer SA. Determining the optimal target blood pressure after thrombectomy: High or low? Neurology. 2017;89:528–529. doi: 10.1212/wnl.0000000000004188

23. Fischer U, Mattle HP. Blood Pressure in Acute Stroke: Still No Answer for Management. Stroke. 2017;48:1717–1719. doi: 10.1161/strokeaha.117.017228

24. Colombari E, Sato MA, Cravo SL, Bergamaschi CT, Campos RR, Jr., Lopes OU. Role of the medulla oblongata in hypertension. Hypertension. 2001;38:549–554. doi: 10.1161/01.hyp.38.3.549

25. Garbin L, Habetswallner F, Clivati A. Vascular reactivity in middle cerebral artery and basilar artery by transcranial Doppler in normals subjects during hypoxia. Ital J Neurol Sci. 1997;18:135–137. doi: 10.1007/bf02048480

26. Maguida G, Shuaib A. Collateral Circulation in Ischemic Stroke: An Updated Review. J Stroke. 2023;25:179–198. doi: 10.5853/jos.2022.02936

27. Powers WJ, Rabinstein AA, Ackerson T, Adeoye OM, Bambakidis NC, Becker K, Biller J, Brown M, Demaerschalk BM, Hoh B, et al. Guidelines for the Early Management of Patients With Acute Ischemic Stroke: 2019 Update to the 2018 Guidelines for the Early Management of Acute Ischemic Stroke: A Guideline for Healthcare Professionals From the American Heart Association/American Stroke Association. Stroke. 2019;50:e344–e418. doi: 10.1161/str.0000000000000211

28. Sandset EC, Anderson CS, Bath PM, Christensen H, Fischer U, Gąsecki D, Lal A, Manning LS, Sacco S, Steiner T, et al. European Stroke Organisation (ESO) guidelines on blood pressure management in acute ischaemic stroke and intracerebral haemorrhage. Eur Stroke J. 2021;6:Ii. doi: 10.1177/23969873211026998

29. Nam HS, Kim YD, Heo J, Lee H, Jung JW, Choi JK, Lee IH, Lim IH, Hong SH, Baik M, et al. Intensive vs Conventional Blood Pressure Lowering After Endovascular Thrombectomy in Acute Ischemic Stroke: The OPTIMAL-BP Randomized Clinical Trial. Jama. 2023;330:832–842. doi: 10.1001/jama.2023.14590

30. Yang P, Song L, Zhang Y, Zhang X, Chen X, Li Y, Sun L, Wan Y, Billot L, Li Q, et al. Intensive blood pressure control after endovascular thrombectomy for acute ischaemic stroke (ENCHANTED2/MT): a multicentre, open-label, blinded-endpoint, randomised controlled trial. Lancet. 2022;400:1585–1596. doi: 10.1016/s0140-6736(22)01882-7

31. Mistry EA, Hart KW, Davis LT, Gao Y, Prestigiacomo CJ, Mittal S, Mehta T, LaFever H, Harker P, Wilson-Perez HE, et al. Blood Pressure Management After Endovascular Therapy for Acute Ischemic Stroke: The BEST-II Randomized Clinical Trial. Jama. 2023;330:821–831. doi: 10.1001/jama.2023.14330

32. Mazighi M, Richard S, Lapergue B, Sibon I, Gory B, Berge J, Consoli A, Labreuche J, Olivot JM, Broderick J, et al. Safety and efficacy of intensive blood pressure lowering after successful endovascular therapy in acute ischaemic stroke (BP-TARGET): a multicentre, open-label, randomised controlled trial. Lancet Neurol. 2021;20:265–274. doi: 10.1016/s1474-4422(20)30483-x

